# Modified bronchoscopy masks mitigate aerosols during gastroscopies

**DOI:** 10.1101/2022.12.06.22283092

**Authors:** Frank Phillips, Jane Crowley, Samantha Warburton, George S.D. Gordon, Adolfo Parra-Blanco

## Abstract

Digestive endoscopy has been proven to produce aerosols (1–3). This represents a risk of infection by COVID-19 and other airborne viruses. A number of protective barriers have been proposed to minimise that risk. Continuous suction of the oral cavity (1), shielding barriers (4,5), masks (6,7), and increasing the distance between patient and endoscopist (8) have been proposed as methods to reduce the exposure of endoscopists and endoscopy staff to aerosols. Here, we present a study that uses modified bronchoscopy masks (Explorer endoscopy facemask, Intersurgical Ltd., United Kingdom) to attenuate aerosol production at the patient’s mouth (bare mask shown in Fig. 1a and in use during an upper GI endoscopy in Fig. 1b). We find that this approach offers 47% (p=0.01) reduction in particle count for particles <5μm in diameter (i.e. aerosols), which are known to spread SARS-CoV-2.

---

To establish the effect of the masks, we measured 13 upper GI endoscopy procedures in which the masks were placed on patients immediately before administering Xylocaine anaesthetic throat spray and were removed after final oral extubation. As a control we measured 33 procedures using normal clinical protocols. We used endoscopy rooms within the same endoscopy suite with room ventilation at 15-17 air changes per hour (measured using a balometer) and similar size, air temperature and humidity levels. Particle counts were measured and analysed using an AeroTrak portable particle counter (TSI, Shoreview MN, model 9500-01) with inlet tube placed 10cm from the patient’s mouth (methodology described in (2)) in order to maximise detection sensitivity and for compatibility with previous studies (1). All present in the room wore enhanced PPE, which minimised the contribution of additional human aerosol sources.

We compared aerosol and droplet concentrations produced from whole procedures (median duration of 7.2 minutes), but we normalise counts to a 20 minute procedure by multiplying total particle count by the appropriate ratio. All statistical analysis was performed using the MATLAB software package (The MathWorks Inc., Massachusetts). Building on existing models of aerosol production in the respiratory tract we use a log-normal distribution to model the distribution of total particle counts (9). For the whole procedure data, a logarithm of the data is first computed, then a t-test is applied to compute p-values. For individual events the data distribution is modelled as the sum of a log-normal and normal distribution to account for negative values of particle counts that can arise from the subtraction step. A Monte-Carlo sampling method is therefore used to provide numerical estimates of p-value and numerically estimate mean ratios and confidence intervals between events (10).

Health Research Authority and ethical approval was granted by the Wales Research Ethics Committee prior to the start of the study (IRAS no. 285595). We included patients undergoing routine upper GI endoscopy on the lists of thirteen different participating endoscopists at the Endoscopy unit of the Nottingham University Hospitals NHS Trust Treatment Centre between October 2020-March 2021. The inclusion criteria were adult patients >18 years with capacity to consent. Biographical data of the patients recorded is shown in Table 1. We found that over the period when the modified bronchoscopy mask was attached the total number of aerosol-sized particles produced was reduced by 47% (95% CI: 16.8% - 65.6%, p<0.01) compared to without masks. We did not find a significant reduction in total particle count for the droplet range (>5µm). Considering individual events, we found that the key aerosol generating events of coughing, extubation and anaesthetic throat spray application were not significantly reduced when the masks were used. This suggests that although the masks are effective at containing continuous low-volume aerosols production, e.g. breathing, they are less effective at containing fast, high-volume production events. We suggest that this is due to the openings in the mask and the constant rate of suctioning: if aerosol production events exceed the suction rate these particles will necessarily escape via these holes.

**Table 1.** Summary table showing demographic data for patients enrolled in this study.

| Mask scenario<br>Variable | No mask on patient | Modified bronchoscopy mask on patient |
| --- | --- | --- |
| *n* | 33 | 12 |
| **Age** | Range: 24-93<br>Median: 63 | Range: 41-83<br>Median: 75.5 |
| **Sex** | Male: 20, Female: 13 | Male: 8, Female: 4 |
| **BMI** | Range: 16.3-38.2<br>Median: 24.8 | Range: 17-38<br>Median: 26.9 |
| **Smoking** | Smoker: 8<br>Non-smoker: 24 | Smoker: 1<br>Non-smoker: 10,<br>Vaper: 1 |
| **Hiatus hernia** | Yes: 9, No: 24 | Yes: 5, No: 7 |
| **Sedation** | Midazolam: 14<br>Throat spray only: 19 | Midazolam: 5<br>Throat spray only: 7 |

**Figure 1.**
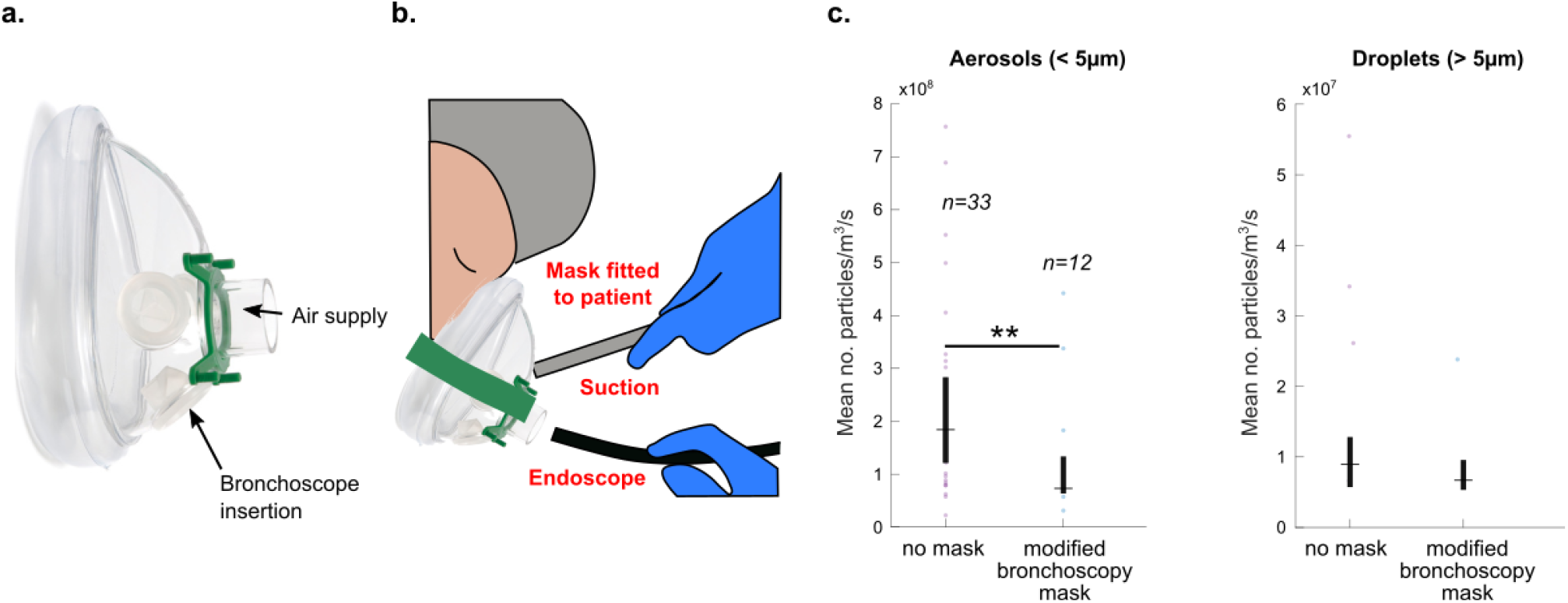
Effect of mitigations on aerosol count. a) Photograph from procedure showing application of modified bronchoscopy mask to patient. b) Effect of modified bronchoscopy masks, showing significant reduction when being used in the <5µm diameter range. *p<0.05, **p<0.01

Overall, we recommend that modified bronchoscopy masks or similar be used to mitigate aerosols during outbreaks of respiratory diseases such as COVID-19. The reduction in particle levels may be sufficient to warrant reduced fallow time as fewer particles means shorter air clearance time, but is not sufficient to remove the need for PPE for healthcare staff. We recommend that improved masks that mitigate aerosols more effectively should be designed.

## Data Availability

Data associated with this publication is available at http://dx.doi.org/10.17639/nott.7112
Code used for data analysis in this publication can be found at https://github.com/gsdgordon/aerosols

## ACKNOWLEDGEMENTS

The authors thank Guru Aithal for critically reviewing the manuscript; Martin James and Bu Hayee for reviewing the study protocol; Matthew Sanderson, Andy Wragg, Nottingham University Hospitals Research and Innovation, University of Nottingham Biomedical Research Centre, Karren Staniforth, Laura Leman, Nina Duffy, Allison Ball and the Endoscopy Unit Staff in their support of the development of this study; the NIHR Aerosol Generating Procedures Group for their support during the study; Tina Rodriguez, Paul Brocklebank, Mirela Pana, Sabina Beg, Stefano Sansone, James Catton, Emilie Wilkes, Lorraine Clark, Andrew Horton, John White, Suresh Vasan Venkatachalapathy, Aida Jawhari, Ioannis Varmpompitis, and Muthuram Rajaram for performing endoscopic procedures in this study.

## PATIENT CONSENT

Obtained

## ETHICS APPROVAL

Wales Research Ethics Committee

